# Human milk feeding and direct breastfeeding improve outcomes for infants with single ventricle congenital heart disease: Propensity score matched analysis of the NPC-QIC registry

**DOI:** 10.1101/2023.04.26.23289126

**Authors:** Kristin M. Elgersma, Julian Wolfson, Jayne A. Fulkerson, Michael K. Georgieff, Wendy S. Looman, Diane L. Spatz, Kavisha M. Shah, Karen Uzark, Anne Chevalier McKechnie

## Abstract

**Background:** Infants with single ventricle (SV) congenital heart disease (CHD) undergo three staged surgeries/interventions, with risk for morbidity and mortality. We estimated the effect of human milk (HM) and direct breastfeeding (BF) on outcomes including necrotizing enterocolitis (NEC), infection-related complications, length of stay (LOS), and mortality.

**Methods:** We analyzed the National Pediatric Cardiology Quality Improvement Collaborative registry (2016–2021), examining HM/BF groups during stage 1 (S1P) and stage 2 (S2P) palliations. We calculated propensity scores for feeding exposures, then fitted Poisson and logistic regression models to compare outcomes between propensity-matched cohorts.

**Results:** Participants included 2491 infants (68 sites). Estimates for all outcomes were better in HM/BF groups. Infants fed exclusive HM before S1P had lower odds of preoperative NEC (OR=0.37, 95% CI=0.17–0.84, p=0.017) and shorter S1P LOS (RR=0.87, 0.78–0.98, p=0.027). During the S1P hospitalization, infants with high HM had lower odds of postoperative NEC (OR=0.28, 0.15–0.50, p<0.001) and sepsis (0.29, 0.13–0.65, p=0.003), and shorter S1P LOS (RR=0.75, 0.66–0.86, p<0.001). At S2P, infants with any HM (0.82, 0.69–0.97, p=0.018) and any BF (0.71, 0.57–0.89, p=0.003) experienced shorter LOS.

**Conclusions:** Infants with SV CHD in high HM and BF groups experienced multiple significantly better outcomes. Given our findings of improved health, strategies to increase the rates of HM/BF in these patients should be implemented. Future research should replicate these findings with granular feeding data and in broader CHD populations, and should examine mechanisms (eg, HM components; microbiome) by which HM/BF benefits these infants.

**Clinical Perspective:** *What is new?:*

- This is the first large, multisite study examining the impact of human milk and breastfeeding on outcomes for infants with single ventricle congenital heart disease.
- All outcome estimates were better in high human milk and breastfeeding groups, with significantly lower odds of necrotizing enterocolitis, sepsis, and infection-related complications; and significantly shorter length of stay at both the neonatal stage 1 palliation and the subsequent stage 2 palliation.
- All estimates of all-cause mortality were substantially lower in human milk and breastfeeding groups, with clinically important estimates of 75%–100% lower odds of mortality in direct breastfeeding groups.

*What are the clinical implications?:*

- There is a critical need for improved, condition-specific lactation support to address the low prevalence of human milk and breastfeeding for infants with single ventricle congenital heart disease.
- Increasing the dose and duration of human milk and direct breastfeeding has strong potential to substantially improve the health outcomes of these vulnerable infants.

## Background

Children born with single ventricle congenital heart disease (SV CHD) are among the most vulnerable of pediatric populations. These infants undergo 3 staged palliative surgeries and/or catheter-based interventions and experience risk for morbidity (eg, necrotizing enterocolitis (NEC)),^1^ developmental delay,^2^ and family maladaptation^3,4^ while incurring the highest hospital costs among United States (US) birth defects.^5^ Mortality rates have been reduced by up to 38% over the past four decades;^6–8^ yet, there remain opportunities to improve outcomes related to development and quality of life.^9^

Feeding for infants with SV CHD is one such developmental area in need of improvement. Human milk (HM) feeding and direct breastfeeding (BF) are agreed upon as the optimal nutrition of choice for hospitalized infants,^10,11^ with a 2023 Science Advisory from the American Heart Association emphasizing that HM and BF are essential to developmental care for infants with critical CHD.^9^ Yet, the SV CHD population has a prevalence of 7% exclusive HM and 9.4% any direct BF at approximately 5 months of age^12^ – far below World Health Organization^10^ and American Academy of Pediatrics^11^ recommendations of exclusive HM feeding for the first 6 months of life, and below the US HealthyPeople 2030 objective of 42.4% exclusive BF through 6 months.^13^ A recent study of lactating parents at 26 US cardiac centers described a lack of institutional and provider support of HM/BF for infants with critical CHD diagnoses including SV CHD.^14^

Inadequate lactation support could be reflective of limited evidence about HM/BF for this population.^9,15^ Human milk and BF positively impact outcomes including NEC,^16^ sepsis,^17^ length of stay,^18^ and mortality^19^ for preterm infants and infants with surgical gastrointestinal anomalies. Little is known, however, about the benefits of HM and BF for infants with CHD. Emerging evidence suggests that exclusive HM feeding both before^20^ and after^21^ neonatal cardiac surgery may be protective against NEC –– a disease with 19–26% mortality in CHD.^22^ However, most studies of HM for infants with CHD are limited by small sample size, heterogenous diagnoses, or risk for statistical bias.^15^ Only one study^21^ has examined HM and outcomes for infants specifically with SV physiology, focusing on the impact of an HM-based fortifier on weight gain. To our knowledge, there is no evidence focused on direct BF and outcomes for infants with CHD of any type. Thus, there is a critical gap in knowledge about HM/BF for infants with SV CHD.

To address this gap in knowledge, we aimed to estimate the effect of HM feeding and direct BF on key outcomes in a large, multisite cohort of infants with SV CHD. We hypothesized that higher dose and/or duration of HM/BF would result in reduced prevalence of NEC, and that we would identify additional benefits related to infection-related complications (including sepsis), time to full feeding volume, hospital length of stay, unplanned hospital readmission, feeding-related hospital readmission, or all-cause mortality.

## Methods

We conducted a propensity score matched cohort analysis of data from the National Pediatric Cardiology Quality Improvement Collaborative (NPC-QIC) registry, which includes infants with SV CHD from >60 US pediatric cardiology centers. Parental informed consent or waiver of consent for registry enrollment was obtained by each NPC-QIC site. For this study, infants with SV CHD who completed S1P were included. General exclusion criteria included family choice not to pursue treatment following birth and S1P admission >3 months of age. The University of Minnesota Institutional Review Board approved this study (STUDY00013371) and deemed it exempt from continuing review. All analyses were conducted using R (versions 4.2.1/4.2.2).^23^ The first author had full access to all data for the study and was responsible for data integrity and analysis.

Our definitions of HM feeding and direct BF, along with NPC-QIC registry feeding measures, have been previously described. Briefly, HM feeding included lactating parent/maternal human milk (MHM) or donor HM via any route, and direct BF was defined as HM directly from a lactating person. We assessed feeding exposures and outcomes at 4 time points (Figure 1). At time points 1 and 2, infants with no S1P preoperative feeding were excluded due to the propensity score assumption of positivity (ie, infants must have had a possibility of receiving the feeding exposure). At time point 3, which is based on S1P discharge data, infants who were never discharged between S1P and S2P were excluded. A flow diagram detailing reasons for inclusion and exclusion at each time point can be found in Figure S1.

**Figure 1.**
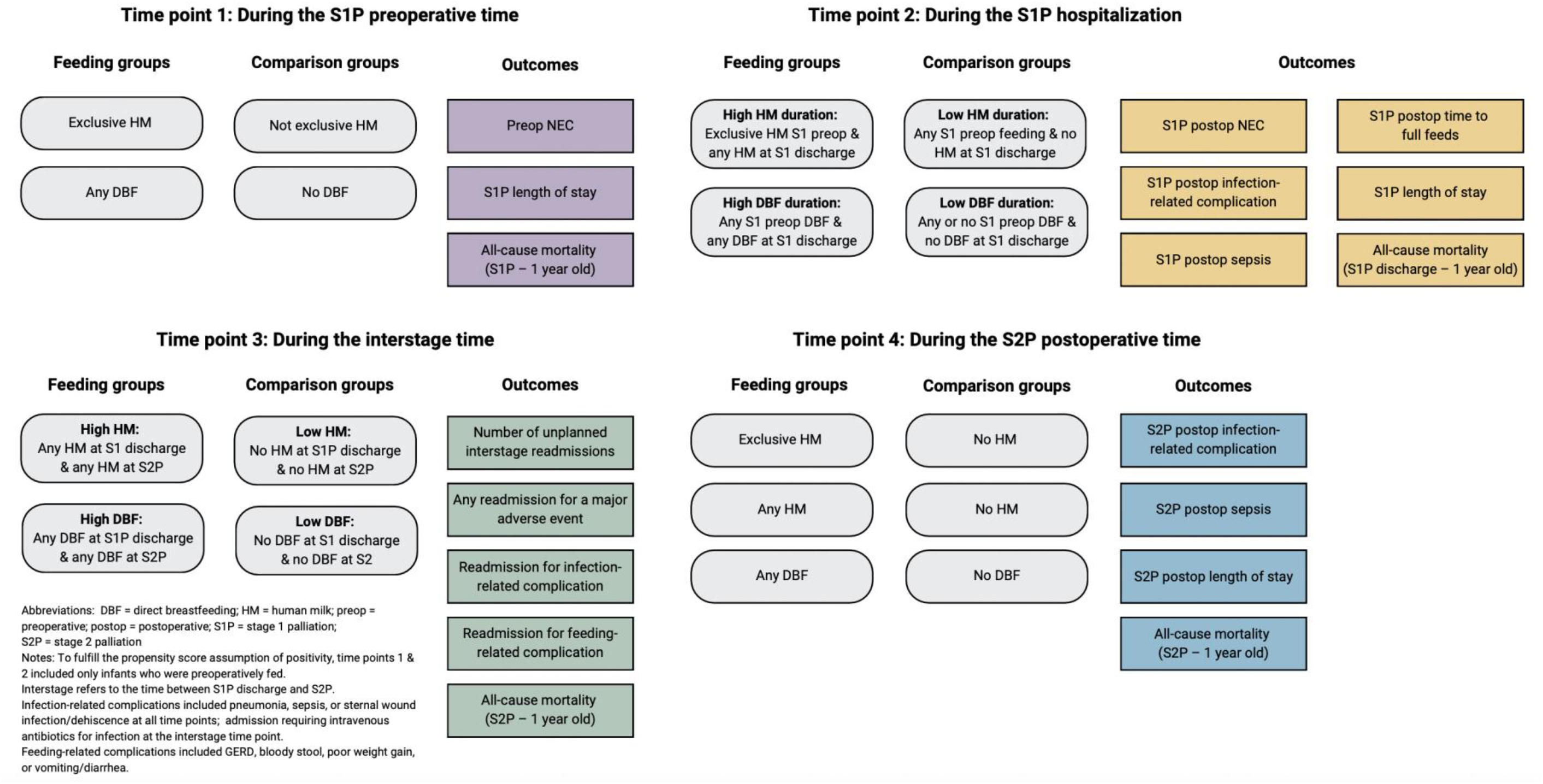
Feeding Groups, Outcomes, and Time Points Examined.

At time point 2, we examined a high HM duration group including infants with exclusive HM preoperatively at S1P who were still receiving any HM at S1P discharge (see Figure 1). These infants were considered to have received HM throughout the course of the S1P hospitalization, in the absence of detailed longitudinal feeding data, and were compared to infants who received any type of preoperative feeding but who were no longer receiving HM at S1P discharge (ie, a low duration group). We examined similar high and low groups for S1P hospitalization duration of direct BF, and high and low HM/BF duration groups across the interstage period.

Outcomes (see Table 1) were selected based on literature from preterm, term, or other neonatal surgical populations demonstrating associations between HM and NEC,^16,17,24–26^ infection,^27^ sepsis,^17,28^ time to full feeds,^29–31^ feeding-related complications,^29,31^ hospital length of stay,^29,30,32^ and all-cause mortality.^17,19^ Our approach to transformation of variables including prematurity, infant race, insurance type, weight-for-age (WAZ) and length-for-age (LAZ) z-scores, and social determinants of health (ie, rural-urban commuting area, median income of residential area, and social deprivation index (SDI))^33^ has been previously described.^12^

**Table 1.** Definitions of Outcomes and Time Points of Outcomes Assessment

| **Outcome** | **Definition** | **Time points** |
| --- | --- | --- |
| Necrotizing enterocolitis | Diagnosed per institution; treated medically or surgically | S1P preop<br>S1P postop |
| S1P length of stay | Date of admission to date of initial discharge | S1P hospitalization |
| S2P postop length of stay | Date of S2P to date of initial discharge | S2P hospitalization |
| Infection-related complication | Diagnosed per institution; aggregate variable including pneumonia, sepsis, or sternal wound infection/dehiscence at all time points; admission requiring IV antibiotics for infection at the interstage time point | S1P postop<br>Interstage<br>S2P postop |
| Sepsis | Diagnosed per institution | S1P postop<br>S2P postop |
| Time to full feeds | Initial postop date on 100 kcal/kg/day enteral feeds | S1P postop |
| Interstage readmission for feeding-related complication | Diagnosed per institution; aggregate variable including GERD, bloody stool, poor weight gain, or vomiting/diarrhea | Interstage<sup>a</sup> |
| Number of unplanned interstage readmissions | Any readmission excluding planned pre-S2P cardiac catheterizations | Interstage<sup>a</sup> |
| Interstage readmission for major adverse event | Any readmission for aspiration, cardiac arrest, infection requiring intravenous antibiotics, cardiac shunt occlusion, life-threatening arrhythmia requiring cardioversion, seizure, or stroke | Interstage<sup>a</sup> |
| All-cause mortality | Death before first birthday; mortality was assessed following each time point of interest | S1P preop<br>Post S1P discharge<br>S2P postop |

Abbreviations: GERD = gastroesophageal reflux disease; IV = intravenous; S1P = stage 1 palliation; S2P = stage 2 palliation; preop = preoperative time; postop = postoperative time
<sup>a</sup>Interstage refers to the time between S1P discharge and S2P

### Analysis

Across the sample, 3.1% of data were missing. We handled missing data via multiple imputation by chained equations (m=20) using the *mice* package^34^ and including all potential covariates, exposure variables, and outcome variables.^34^

We conducted propensity score matched analyses using the *MatchThem* package^35^ for imputed data to determine the average treatment effect among the treated (ATT). Propensity score matching supports causal inference with reduced bias and is particularly useful in HM/BF research, as true randomization is not possible. Propensity score models (ie, logistic regression models for the probability of being in the feeding group of interest) were created for each exposure/outcome combination (Table S1). As recommended by Brookhart et al.,^36^ variables strongly related to the outcome were included in these models, while variables related to the exposure but not the outcome were excluded.

Propensity scores were calculated within each imputed data set, using nearest neighbor matching with a caliper ranging from 0.2–0.3 of the standard deviation of the logit of the propensity score. Matching ratios of up to 10:1 and matching with replacement were explored, with limits on the number of times a control could be reused. Covariate balance was assessed using the *cobalt* package^37^ to obtain absolute standardized mean differences (SMD; ie, the largest SMD for each covariate across imputations^35^), with an SMD <0.10 considered to be acceptably balanced. We considered interaction and polynomial terms to aid in covariate balance, and those covariates that could not be balanced were included in the final outcome regression models. Further details of propensity score models are found in Table S1, and covariate balance visualized in Figures S2 and S3.

We then fitted Poisson or logistic regression models to compare outcomes between propensity matched cohorts using the “svlglm” function from the *survey* package^38^ for robust standard error calculation,^35^ with Rubin’s^39^ rules used to obtain pooled odds ratios (OR) or rate ratios (RR) and 95% confidence intervals (CI). Poisson regression was used to model the outcomes of hospital length of stay, time to full feeds, and the number of unplanned interstage readmissions, with additional models fitted for these outcomes using a Gaussian distribution for comparison. Logistic regression models were fitted for the remaining binary outcomes. For rare binary outcomes resulting in issues with separation (eg, 0% outcome prevalence in one feeding group), logistic regression models were refitted with mean bias-reducing score adjustment^40,41^ using the “brglmFit” function in the *brglm2* package.^42^ Sensitivity analysis included inverse probability weighting using the propensity score to calculate the ATT on the same imputed data.

Covariate selection for the analyses examining HM or BF during the S1P hospitalization (Figure 1, time point 2) was challenging, as we were unable to determine dates of, for example, major postoperative procedures or complications. Therefore, we were liberal in including potentially important covariates in propensity score models for the main analyses and also conducted sensitivity analyses using propensity score models with more limited, baseline covariates.

## Results

Of 2697 infants in the NPC-QIC registry, 2491 from 68 sites met general study inclusion criteria. Of note, all infants diagnosed with S1P preoperative NEC completed S1P and were included in the study. Key sample characteristics and sample-wide outcomes can be found in Table 2.

**Table 2.** Characteristics and Outcomes of Interest Among the Full National Pediatric Cardiology Quality Improvement Project Registry Sample (N = 2491)

| Sample characteristics: mean (SD) or n (%) |  | Outcomes of interest: mean (SD) or n (%) |  |
| --- | --- | --- | --- |
| Sex |  | S1P preop NEC |  |
| Female | 988 (39.7) | Yes | 58 (2.3) |
| Male | 1501 (60.3) | No | 2433 (97.7) |
| Ambiguous or Unknown | 2 (0.1) | S1P postop NEC |  |
| Race |  | Yes | 324 (13.0) |
| Another Race/Multi-Race | 297 (12.3) | No | 2167 (87.0) |
| Black/African American | 397 (16.4) | S1P postop infection-related complication<sup>a</sup> |  |
| White | 1721 (71.3) | Yes | 386 (15.5) |
| Unknown | 76 | No | 2105 (84.5) |
| Hispanic or Latino/a Ethnicity |  | S1P postop sepsis |  |
| Yes | 390 (16.4) | Yes | 98 (5.1) |
| No | 1987 (83.6) | No | 1842 (94.9) |
| Unknown | 114 | Unknown or NA | 551 |
| Insurance type |  | S1P postop time to full feeds (d) | 16 (17) |
| Government | 1302 (54.6) | Unknown or NA | 254 |
| Private/Self | 1084 (45.4) | S1P length of stay (d) | 48 (32) |
| Unknown | 105 | Unknown or NA | 550 |
| Median income of residential ZCTA | 65,667 (23,849) | # of unplanned interstage readmissions | 1.92 (1.27) |
| Unknown | 69 | Unknown or NA | 1527 |
| SDI score of residential ZCTA | 51 (28) | Interstage readmission for major adverse event<sup>b</sup> |  |
| Unknown | 63 | Yes | 67 (7.0) |
| Rural-urban commuting area |  | No | 897 (93.0) |
| Metropolitan | 1971 (81.1) | Unknown or NA | 1527 |
| Micropolitan | 254 (10.5) | Interstage readmission for infection-related complication<sup>a</sup> |  |
| Rural | 204 (8.4) | Yes | 136 (14.1) |
| Unknown | 62 | No | 828 (85.9) |
| Preterm (<37 weeks) |  | Unknown or NA | 1527 |
| Yes | 307 (12.4) | Interstage readmission for feeding-related complication<sup>c</sup> |  |
| No | 2161 (87.6) | Yes | 357 (37.0) |
| Unknown | 23 | No | 607 (63.0) |
| WAZ at birth | -0.18 (1.10) | Unknown or NA | 1527 |
| Unknown | 73 | S2P postop infection-related complication<sup>a</sup> |  |
| Primary cardiac diagnosis |  | Yes | 107 (5.8) |
| HLHS | 1757 (70.5) | No | 1739 (94.2) |
| Other SV | 734 (29.5) | Unknown or NA | 645 |
| Secondary cardiac diagnosis |  | S2P postop sepsis |  |
| Ascending aorta restriction | 220 (8.8) | Yes | 62 (3.4) |
| Restrictive/intact atrial septum | 1378 (55.3) | No | 1779 (96.6) |
| Other | 640 (25.7) | Unknown or NA | 650 |
| None | 253 (10.2) | S2P postop LOS | 19 (24) |
| Other major anomaly |  | Unknown or NA | 758 |
| Yes | 190 (7.6) | All-cause mortality<sup>d</sup> |  |
| No | 2301 (92.4) | Yes | 357 (14.3) |
| Major genetic syndrome |  | No | 2134 (85.7) |
| Yes | 314 (12.6) | Abbreviations: d = days; HLHS = hypoplastic left heart syndrome; LOS = length of stay; NA = not applicable; NEC = necrotizing enterocolitis, postop = postoperative; preop = preoperative; S1P = stage 1 palliation; S2P = stage 2 palliation, SV = single ventricle; ZCTA = zip code tabulation area<br><sup>a</sup>Includes pneumonia, sepsis, wound infection/dehiscence, and (in interstage) infection requiring intravenous antibiotics<br><sup>b</sup>Includes aspiration, cardiac arrest, infection requiring intravenous antibiotics, cardiac shunt occlusion, life-threatening arrhythmia requiring cardioversion, seizure, and stroke<br><sup>c</sup>Includes gastroesophageal reflux disease, bloody stool, poor weight gain, and vomiting/diarrhea<br><sup>d</sup>Between stage 1 palliation and 1 year of age |  |
| No | 2177 (87.4) |  |  |
| Age at S1P admission | 1.13 (4.87) |  |  |
| Comprehensive parental postnatal support |  |  |  |
| Yes | 2360 (94.7) |  |  |
| No | 131 (5.3) |  |  |
| S1P preoperative enteral feeding |  |  |  |
| Yes | 1440 (57.8) |  |  |
| No | 1051 (42.2) |  |  |
| Discharged after S1 |  |  |  |
| Yes | 2205 (89.5) |  |  |
| No | 259 (10.5) |  |  |
| Unknown or NA | 27 |  |  |

### Propensity score matched analyses

**S1P hospitalization.** The estimates for all S1P hospitalization outcomes were better in the high HM and BF groups, although not all reached statistical significance (see Tables 3 and 4). Infants with SV CHD who were preoperatively fed exclusive HM had 63% lower odds of preoperative NEC (OR=0.37, 95% CI=0.17–0.84, p=0.017) and a 13% reduction in mean S1P length of stay (RR=0.87, 0.78–0.98, p=0.027).

**Table 3.** Average Treatment Effect Among the Treated of Exclusive Human Milk Feeding and Any Direct Breastfeeding During the Stage 1 Palliation Preoperative Time for Key Outcomes in Propensity Score Matched Cohorts

| **Exclusive HM (n=934) vs. Not exclusive HM (n=331)** |  |  |  |
| --- | --- | --- | --- |
|  | **OR or RR** | **95% CI** | **p value** |
| Outcome |  |  |  |
| Preop NEC | 0.37<sup>a</sup> | (0.17–0.84) | **0.017** |
| S1P hospital LOS | 0.87<sup>b</sup> | (0.78–0.98) | **0.027** |
| All-cause mortality<sup>c</sup> | 0.70<sup>a</sup> | (0.46–1.07) | 0.099 |
| **Any DBF (n=378) vs. No DBF (n=920)** |  |  |  |
|  | **OR or RR** | **95% CI** | **p value** |
| Outcome |  |  |  |
| Preop NEC | 0.73<sup>a</sup> | (0.25–2.12) | 0.566 |
| S1P hospital LOS | 0.94<sup>b</sup> | (0.83–1.07) | 0.361 |
| All-cause mortality<sup>c</sup> | 0.73<sup>a</sup> | (0.44–1.21) | 0.227 |

Abbreviations: CI = confidence interval; DBF = direct breastfeeding; HM = human milk; LOS = length of stay; NEC = necrotizing enterocolitis; OR = odds ratio; preop = preoperative; RR = rate ratio
<sup>a</sup>Analysis included logistic regression; estimate presented as odds ratio
<sup>b</sup>Analysis included Poisson regression; estimate presented as rate ratio
<sup>c</sup>Between stage 1 palliation and 1 year of age

**Table 4.** Average Treatment Effect Among the Treated of High Human Milk Feeding or Direct Breastfeeding Duration in the Stage 1 Palliation Hospitalization for Key Outcomes in Propensity Score Matched Cohorts

| **High HM duration: Exclusive preop HM + any HM at discharge (n=603) vs.**<br>**Low HM duration: Any type of preop feeding but no HM at discharge (n=464)** |  |  |  |
| --- | --- | --- | --- |
| Outcome | OR or RR | 95% CI | p value |
| Postop NEC | 0.28<sup>a</sup> | (0.15–0.50) | **<0.001** |
| Infection-related postop complication<sup>c</sup> | 0.48<sup>a</sup> | (0.25–0.91) | **0.025** |
| Postop sepsis | 0.29<sup>a</sup> | (0.13–0.65) | **0.003** |
| Time to full feeds | 0.95<sup>b</sup> | (0.82–1.10) | 0.492 |
| S1P hospital LOS | 0.75<sup>b</sup> | (0.66–0.86) | **<0.001** |
| All-cause mortality<sup>d</sup> | 0.54<sup>a</sup> | (0.20–1.46) | 0.226 |

**High DBF duration: Any preop DBF + any DBF at discharge (n=102) vs. Low DBF duration: Any type of preop feeding but no DBF at discharge (n=1004)**
| Outcome | OR or RR | 95% CI | p value |
| --- | --- | --- | --- |
| Postop NEC | 0.67<sup>a</sup> | (0.29–1.56) | 0.355 |
| Infection-related postop complication<sup>c</sup> | 0.66<sup>a</sup> | (0.20–2.21) | 0.501 |
| Postop sepsis | 0.00<sup>a</sup> | NC<sup>e</sup> | **<0.001** |
| Time to full feeds | 0.89<sup>b</sup> | (0.71–1.12) | 0.317 |
| S1P hospital LOS | 0.77<sup>b</sup> | (0.66–0.90) | **0.001** |
| All-cause mortality<sup>d</sup> | 0.25<sup>a</sup> | (0.03–2.29) | 0.218 |

Abbreviations: CI = confidence interval; DBF = direct breastfeeding; HM = human milk; LOS = length of stay; NEC = necrotizing enterocolitis; OR = odds ratio; postop = postoperative; preop = preoperative; RR = rate ratio; S1P = stage 1 palliation
<sup>a</sup>Analysis included logistic regression; estimate presented as odds ratio
<sup>b</sup>Analysis included Poisson regression; estimate presented as rate ratio
<sup>c</sup>Includes postoperative pneumonia, sepsis, and wound infection
<sup>d</sup>Between stage 1 palliation and 1 year of age
<sup>e</sup>Due to rare occurrence of the outcome, confidence intervals were not computable

Infants with high HM feeding duration across the S1P hospitalization had 72% lower odds of postoperative NEC (0.28, 0.15–0.50, p<0.001), 52% lower odds of an infection-related postoperative complication (0.48, 0.25–0.91, p=0.025), 71% lower odds of postoperative sepsis (0.29, 0.13–0.65, p=0.003), and a 25% reduction in S1P length of stay (RR=0.75, 0.66–0.86, p<0.001).

Infants with high direct BF duration across the S1P hospitalization had 100% lower odds of postoperative sepsis in the main analysis, with a similar result in the bias-corrected sensitivity analysis (OR=0.07, 0.02–0.22, p<0.001). In the unmatched cohort, this finding corresponded to 0% vs. 6.6% prevalence of S1P postoperative sepsis in the high vs. low BF duration groups. Infants with high BF duration also had a 23% reduction in S1P length of stay (0.77, 0.66–0.90, p=0.001).

#### Interstage and S2P hospitalization

The results of the propensity score matched analyses for the interstage and S2P hospitalization time points are in Tables 5 and 6. Again, the estimates for all outcomes were slightly-to-substantially better in the high HM and BF groups. Infants with high interstage BF duration had 100% lower odds of mortality between S2P and 1 year old in the main analysis, corresponding to 0% vs. 3.3% prevalence in high vs. low BF groups in the unmatched sample and 77% reduced odds in the bias-corrected analysis (0.23, 0.09–0.58, p=0.002).

**Table 5.** Average Treatment Effect Among the Treated of High Interstage Human Milk Feeding or Direct Breastfeeding Duration (Stage 1 Palliation Discharge to Stage 2 Palliation) for Key Outcomes in Propensity Score Matched Cohorts

| Outcome | OR or RR | 95% CI | p value |
| --- | --- | --- | --- |
| # of unplanned interstage readmissions | 0.97<sup>a</sup> | (0.72–1.30) | 0.835 |
| Any interstage readmission for adverse events<sup>c</sup> | 0.89<sup>b</sup> | (0.52–1.54) | 0.684 |
| Infection-related interstage readmission<sup>d</sup> | 0.86<sup>b</sup> | (0.30–2.45) | 0.684 |
| Feeding-related interstage readmission<sup>e</sup> | 0.66<sup>b</sup> | (0.30–1.47) | 0.310 |
| All-cause mortality<sup>f</sup> | 0.31<sup>b</sup> | (0.04–2.63) | 0.284 |

| Outcome | OR or RR | 95% CI | p value |
| --- | --- | --- | --- |
| # of unplanned interstage readmissions | 0.89<sup>a</sup> | (0.61–1.30) | 0.531 |
| Any interstage readmission for adverse events<sup>c</sup> | 0.97<sup>b</sup> | (0.54–1.74) | 0.911 |
| Infection-related interstage readmission<sup>d</sup> | 0.97<sup>b</sup> | (0.32–2.98) | 0.959 |
| Feeding-related interstage readmission<sup>e</sup> | 0.46<sup>b</sup> | (0.20–1.06) | 0.069 |
| All-cause mortality<sup>f</sup> | 0.00<sup>b</sup> | NC<sup>g</sup> | <0.001 |

Abbreviations: CI = confidence interval; DBF = direct breastfeeding; HM = human milk; NC = not computable; OR = odds ratio; RR = rate ratio; S1P = stage 1 palliation; S2P = stage 2 palliation
<sup>a</sup>Analysis included Poisson regression; estimate presented as rate ratio
<sup>b</sup>Analysis included logistic regression; estimate presented as odds ratio
<sup>c</sup>Includes aspiration, cardiac arrest, infection requiring intravenous antibiotics, cardiac shunt occlusion, life-threatening arrhythmia requiring cardioversion, seizure, and stroke
<sup>d</sup>Includes pneumonia, sepsis, wound infection/dehiscence, and infection requiring intravenous antibiotics
<sup>e</sup>Includes gastroesophageal reflux disease, bloody stool, poor weight gain, and vomiting/diarrhea
<sup>f</sup>Between stage 2 palliation and 1 year of age
<sup>g</sup>Due to rare occurrence of the outcome, confidence intervals were not computable

**Table 6.** Average Treatment Effect Among the Treated of Human Milk Feeding and Direct Breastfeeding at Stage 2 Palliation for Key Outcomes in Propensity Score Matched Cohorts

| **Any HM at S2P (n=785) vs. No HM at S2P (n=1062)** |  |  |  |
| --- | --- | --- | --- |
|  | **OR or RR** | **95% CI** | **p value** |
| Outcome |  |  |  |
| Infection-related S2P postop complication<sup>c</sup> | 0.94<sup>a</sup> | (0.53–1.68) | 0.838 |
| S2P postop sepsis | 0.61<sup>a</sup> | (0.28–1.30) | 0.196 |
| S2P postop hospital LOS | 0.82<sup>b</sup> | (0.69–0.97) | **0.018** |
| All-cause mortality<sup>d</sup> | 0.85<sup>a</sup> | (0.41–1.76) | 0.661 |
| **Exclusive HM at S2P (n=130) vs. No HM at S2P (n=947)** |  |  |  |
|  | **OR or RR** | **95% CI** | **p value** |
| Outcome |  |  |  |
| Infection-related S2P postop complication<sup>c</sup> | 0.56<sup>a</sup> | (0.16–1.92) | 0.353 |
| S2P postop sepsis | 0.49<sup>a</sup> | (0.12–1.98) | 0.315 |
| S2P postop hospital LOS | 0.75<sup>b</sup> | (0.57–0.99) | **0.040** |
| All-cause mortality<sup>d</sup> | 0.49<sup>a</sup> | (0.18–1.35) | 0.166 |
| **Any DBF at S2P (n=173) vs. No DBF at S2P (n=1674)** |  |  |  |
|  | **OR or RR** | **95% CI** | **p value** |
| Outcome |  |  |  |
| Infection-related S2P postop complication<sup>c</sup> | 0.39<sup>a</sup> | (0.11–1.34) | 0.136 |
| S2P postop sepsis | 0.87<sup>a</sup> | (0.18–4.12) | 0.861 |
| S2P hospital LOS | 0.71<sup>b</sup> | (0.57–0.89) | **0.003** |
| All-cause mortality<sup>d</sup> | 0.24<sup>a</sup> | (0.03–1.88) | 0.174 |

Abbreviations: CI = confidence interval; DBF = direct breastfeeding; HM = human milk; LOS = length of stay; NEC = necrotizing enterocolitis; postop = postoperative; OR = odds ratio; RR = rate ratio; S2P = stage 2 palliation
<sup>a</sup>Analysis included logistic regression; estimate presented as odds ratio
<sup>b</sup>Analysis included Poisson regression; estimate presented as rate ratio
<sup>c</sup>Includes postoperative pneumonia, sepsis, and wound infection
<sup>d</sup>Between stage 2 palliation and 1 year of age

At S2P, all HM/BF groups had a significant reduction in postoperative hospital length of stay with mean reductions of 18% (0.82, 0.69–0.97, p=0.018) for any HM; 25% (0.75, 0.57–0.99, p=0.040) for exclusive HM, and 29% (0.71, 0.57–0.89, p=0.003) for any BF.

### Sensitivity analyses

The results of sensitivity analyses using inverse probability weighting; limited baseline covariates for S1P hospitalization propensity score models; and Gaussian distribution for hospital length of stay, time to full feeds, and interstage readmissions supported the main conclusions of the above analyses, with estimates that were similar in direction and magnitude (data not shown; available upon request from the authors).

## Discussion

This study addresses a critical gap in knowledge as the first large, multisite analysis of the relationship between HM or direct BF and several key outcomes for infants with SV CHD. In our propensity score matched cohorts, all outcome estimates at four time points during the first year of life were better in HM/BF groups, with many results reaching statistical significance and substantial clinical significance across the board. We will focus our discussion on results relating to NEC, sepsis and infection, length of stay, and mortality.

### Necrotizing enterocolitis

We found that infants with higher exposure to HM feeding had lower odds of S1P preoperative and postoperative NEC. These findings are consistent with two decades of research in preterm populations, and are important in light of a 2022 review by Burge et al.^43^ outlining potential differences between NEC in preterm infants and the cardiac NEC experienced by infants with CHD. Burge and colleagues suggested that cardiac NEC is, in part, a function of impaired gut perfusion, with resulting hypoperfusion and mesenteric ischemia contributing to an endothelial inflammatory response with associated gut permeability and pathogenic translocation. Dysbiosis of the gut microbiome related to high systemic inflammation,^44^ prophylactic antibiotics, and delayed enteral feeding are known to play a role in NEC and intestinal injury in neonates.^43^

Despite potential differences in etiology between preterm and cardiac NEC, our findings suggest that the protective benefits of HM demonstrated for preterm infants extrapolate to the SV CHD population. Four recent systematic reviews and meta-analyses^16,16,24,26^ demonstrate convincing reductions in preterm NEC due to provision of HM and/or avoidance of infant formula (eg, 68% reduced risk;^25^ 4% lower incidence^16^). Few previous studies, however, have examined the relationship between HM and NEC in infants with CHD.^20,21,45^ Our results align with Cognata et al.’s^20^ well-designed retrospective cohort study which reported 83% lower odds of preoperative NEC for exclusive HM-fed infants with critical CHD.

In another study conducted at the same institution as in Cognata et al., with the same population,^20^ Kataria-Hale et al.^45^ found no difference in postoperative NEC related to exclusive preoperative HM. The authors hypothesized that postoperative feeding practices at the time of NEC development may have been more influential. Our findings lend support to this hypothesis, as infants with high HM feeding during the S1P hospitalization had 72% lower odds of postoperative NEC. This result is particularly intriguing, as the literature regarding critical CHD has often focused on preoperative NEC due to controversy about the safety of preoperative enteral feeding. However, the prevalence of postoperative NEC, as diagnosed per institution, was higher in our sample (ie, 13.0% postoperatively vs. 2.3% preoperatively), emphasizing the need to reduce NEC throughout the entire S1P hospitalization. We also recommend further examination of the type, timing, and delivery mode of milk fortification as standard postoperative protocol. While Blanco et al.’s^21^ 2022 randomized controlled trial (RCT) testing an HM-based fortifier suggests that an exclusive HM diet may reduce the incidence of postoperative NEC for infants with SV CHD, this fortifier is not yet available in the US and the study was not powered for the NEC outcome. Exposure to bovine-milk-based fortifier/formula, which has been shown to increase the risk of NEC,^46^ is the current standard of care. Future research is needed to identify ways to increase the dose and duration of postoperative HM while supporting growth and development, with exclusive HM feeding a potentially critical intervention to reduce NEC-related morbidity and mortality for infants with SV CHD.

#### Potential mechanisms

In recent years, there has been increased focus on the cellular, molecular, and nutritional composition of HM and on the relationship between HM and the infant gut microbiome-immune axis. Research has elucidated mechanisms influencing the development of NEC in preterm infants, with HM components such as HM oligosaccharides,^47–50^ exosomes,^51,52^ fatty acids and lipids,^53^ lactoferrin,^54^ immunoglobulins,^54,55^ and many other bioactive factors^56^ offering tailored protection against NEC and other hospital-associated diseases. Emerging evidence reveals that HM from the infant’s own lactating parent (ie, MHM) is associated with epigenetic variation in DNA methylation,^57,58^ which may provide protection against oxidative stress that could contribute to NEC. These HM components are closely related to healthy development of the infant gut microbiome.^59^ Studies have characterized the gastrointestinal microbiome^60^ of preterm populations, revealing frequent dysbiosis driven by exposure to infant formula, antibiotics, and delivery mode (ie, cesarean section) that could contribute to NEC.^61^ Interestingly, two 2022 RCTs^62,63^ examining infant fortifiers highlight the crucial role of MHM in positively shaping the preterm gastrointestinal microbiome, with Kumbhare et al.^62^ identifying volume of MHM as the strongest predictor of the preterm infant’s gut microbiota.

Knowledge about the gut microbiome of infants with CHD is only beginning to emerge,^64–69^ and there has been no investigation into HM composition in the context of CHD. A 2022 study by Huang et al.^64^ provides the first comprehensive evidence on the gut microbiome of neonates with critical CHD, reporting dysbiosis characterized by increased pathogens (eg, *Enterococcaceae*, *Enterobacteriaceae*) and decreased beneficial organisms (eg, *Bifidobacterium*, *Lactobacillus*) – a profile that shares similarities with the gut microbiome of very-low-birth-weight infants^60^ – with consequent inflammatory and immune imbalances potentially contributing to poor clinical outcomes, including NEC. Huang and colleagues note the key role of HM/BF (eg, human milk oligosaccharides) in establishing normal gut *Bifidobacterium* colonization and reducing pathogenic activity, and speculate that low HM/BF prevalence could contribute to gut dysbiosis in the context of CHD.^64^ The authors propose *Bifidobacterium* and oligosaccharide supplementation for infants with critical CHD, but stop short of recommending improved lactation support for these infants and their families as a mechanism to promote intestinal homeostasis. Of the remaining abstracts,^69^ studies,^65,67,68^ or reviews^66^ identified on the topic of the gut microbiome in patients with CHD, only one briefly mentions HM/BF^68^ and none discuss HM/BF as a therapeutic intervention for gut dysbiosis.

The omission of HM/BF from the CHD gut microbiome literature is not entirely surprising as support for HM/BF in the CHD population has been historically inadequate and under prioritized by the healthcare team,^14,70–72^ contributing to extremely low prevalence of these feeding practices.^12^ We also speculate that, as the underlying cause of cardiac NEC may be different than in preterm infants, some providers might assume that HM is not similarly protective for infants with CHD. Both Huang et al.’s^64^ novel research demonstrating similarities between the gut microbiome of preterm infants and those with critical CHD and our findings of strong associations between HM and reduced NEC would discount this assumption. Furthermore, in animal studies focused on the mechanistic relationship between HM and preterm NEC, a common method of NEC–induction involves subjecting mice to hypoxia, infant formula, and introduction of lipopolysaccharide to induce inflammation^73^ – a process with clear analogies to the SV CHD clinical course. Infants with SV CHD and their lactating parents also experience many of the same risk factors for gut dysbiosis as in preterm birth (eg, antibiotic use, delayed fetal development, formula supplementation, lack of direct BF, breast pump use).^60,74,75^ Therefore, there is a critical need for CHD researchers and clinicians to learn from and build upon the foundation of lactation research in preterm populations, with the relationship between HM components/microbiota and infant gut microbiome alterations in the context of critical CHD an important area for future study.

### Sepsis and infection

Infants with high HM feeding duration in the S1P hospitalization had lower odds of postoperative sepsis and infection-related complications, while those with high direct BF duration had 100% lower odds of postoperative sepsis. No infants in the interstage high BF group were readmitted for sepsis, although the overall prevalence of interstage sepsis was low. HM has been associated with lower rates of sepsis^16,17,19,30,76,77^ and infection^32^ for preterm infants and infants with surgical gastrointestinal anomalies, with protective mechanisms likely similar to those previously described for NEC (eg, reduced gut dysbiosis with subsequent lower risk of pathogenic gut bacteria translocation^78^). A 2023 study by Ghosh et al.^79^ reported a 2.58 times increase in the odds of postoperative infection (ie, bloodstream infection, surgical site infection, ventilator-associated pneumonia; p = 0.040) associated with exclusive infant formula, compared to exclusive HM, for infants undergoing cardiac surgery at a single center in India. This study, however, is limited in that infants in the exclusive HM group were substantially older (ie, median 60 days vs. 15 days) and underwent less complicated procedures, on average. Furthermore, Ghosh and colleagues described COVID-19-related restrictions on maternal bedside presence that may have disproportionately affected infants living in remote areas or families who could not travel, and socioeconomic factors or other social determinants of health were not reported. We identified no studies specifically examining the relationship between direct BF and infection or sepsis in any hospitalized neonates.

Our finding of reduced odds of sepsis in high BF groups offers novel evidence that direct BF as a mode of HM delivery may be particularly beneficial in preventing sepsis in infants with SV CHD. Interestingly, a 2019 study^75^ using a robust, multi-method analytical approach identified mode of feeding as a key contributor to the HM microbiome, with HM fed directly from the breast exhibiting significantly decreased pathogenic *Enterobacteriaceae* and *Enterococcaceae*, high beneficial *Bifidobacterium*, and increased microbial richness and diversity compared to expressed HM. There is also emerging evidence supporting a retrograde inoculation hypothesis, in which the flow of milk from an infant’s oral cavity back into the mammary ductal system shapes the microbiome of both the infant and the lactating parent.^75,80–83^ This microbial communication between infant and parent during BF could be one mechanism to explain the changes in the immunological composition of HM in response to pathogenic organisms that can lead to sepsis^84^ and suggests that direct BF could confer critical protection to vulnerable infants, including those with SV CHD.

### Length of stay

Hospital length of stay was consistently lower in the HM and BF groups at all time points examined. Interestingly, infants with exclusive HM preoperatively at S1P had a 13% mean reduction in S1P length of stay. The preoperative time typically lasts less than a week and is followed by high-risk intervention, an often complicated recovery, and a lengthy hospital stay (mean 48 ± 32 days). It is notable that this short exposure to exclusive HM appeared to have lasting impact in our matched cohort.

Similarly, infants with high HM feeding duration in the S1P hospitalization had a 25% lower mean S1P length of stay, and those with high BF duration had a 23% lower mean S1P length of stay. Associations between HM and shorter length of stay have been demonstrated for preterm infants^25,85^ and infants with other surgical anomalies,^18,30,77,86^ although results are inconsistent, may be dose dependent,^18,30,86^ and may differ between MHM^85^ and donor HM.^25^ To our knowledge, only one study has examined associations between HM and length of stay for infants with CHD. Yu et al.^87^ found that infants fed HM had a 3.9 days shorter mean length of stay compared to a formula feeding group in a cohort with varied CHD diagnoses; however, this study exhibits high risk of bias.^15^

We did not identify any studies examining direct BF and length of stay for infants with surgical congenital anomalies. The preterm literature similarly focuses primarily on HM feeding as a nutritional entity rather than on the mode of feeding, although Suberi et al.^88^ reported an association between direct BF as the first mode of oral feeding (compared to bottle) for preterm infants and ∼1 week earlier NICU discharge. Our study is unique in that it addresses the critical gap in knowledge about direct BF in this population and suggests some differential benefits.

Establishing causality between HM or BF and S1P length of stay in this population is challenging. Hospital length of stay has often been considered as a predictor of HM/BF practices rather than as an outcome impacted by infant feeding,^89^ and the psychological stress of extended hospitalization, postoperative complications, or family/work obligations could impact a parent’s ability to provide HM/BF. These are potentially unmeasured confounders that could impact our results, and there is likely some element of multidirectionality between HM/BF and hospital length of stay. An individual infants’ hospital stay can be extended for many reasons, and numerous potential disruptions to feeding development have been outlined by Jones et al.^90^ However, in light of the limitations of the registry data in this study, our propensity score models included multiple indicators of a complicated clinical course (Table S1; eg, preoperative instability, intubation duration, major postoperative procedures) and parental access to supportive resources (eg, insurance type, SDI score of ZCTA, comprehensive parental support delivered), along with clinical site to account for differences in institutional practices, protocols, and level of lactation support. Even when accounting for these potential confounding variables, associations between HM or BF and length of stay remained strong. As a partial explanation, we hypothesize a causal pathway between HM/BF; reduced incidence of NEC, infection, and sepsis; and S1P hospital length of stay.^29^

The potential for a causal effect of HM and/or BF on length of stay is further supported by the results from the S2P hospitalization, in which the temporal relationship between HM/BF and S2P postoperative length of stay was more clearly defined. Once again, length of stay was significantly shorter in all HM/BF groups, with reductions similar in magnitude to those in the S1P analysis (ie, 18%–29%). Multisite, prospective, longitudinal studies with granular feeding data are needed to confirm these results, and future research should also elucidate potential mechanistic causes. Within-site variation, by individual providers,^91^ of practices that impact both HM/BF and length of stay could also be important for future research and potential practice modification. Given that hospital length of stay has been identified as the key driver of hospital costs for infants with CHD,^92^ our findings suggest that improving HM/BF prevalence has potential to not only improve the health of infants with SV CHD, but also to reduce economic costs for families, payers, and institutions.

### Mortality

While most analyses of all-cause mortality did not reach statistical significance in this study, all estimates were substantially lower in the HM and BF groups. The clinical importance was particularly striking for direct BF groups, with estimates of 75%–100% lower odds of mortality. Previous research in preterm^17,19^ and other neonatal surgical populations^18^ suggests an association between HM and mortality, and our study provides initial evidence of a potential difference in survival related to HM/BF practices for infants with SV CHD. Regarding direct BF, it may be tempting to assume that infants who are able to BF are less “sick” than those who are not and therefore at lower risk for death; however, the propensity score models for our cohort included many indicators of an infant’s relative sickness and previous research has described successful direct BF in the context of severe CHD disease presentation.^14^ We speculate that the achievement of the complex neurodevelopmental skill of direct BF could both reflect and promote improved clinical status. Future studies including detailed data on feeding method/dose and reasons for infant death are needed to support more robust survival analyses in this population.

### Limitations

Limitations of our study include those inherent in analysis of multisite registries, such as potential for inaccurate, inconsistent, or missing data. While registry data in this rare disease population offers advantages, we could not fully characterize an infant’s HM/BF trajectory over time, and the dose could have varied widely within groups. We also did not have information on the timing of outcomes (eg, dates of NEC or sepsis diagnosis). Therefore, although our analytical approach was designed to reduce bias, results should be interpreted as hypothesis-generating rather than confirmatory. For infants in HM groups, we were unable to determine whether the HM was from the lactating parent or donor HM. It is increasingly clear that MHM and donor HM are not equivalent, as many of the protective bioactive components of MHM are eliminated during pasteurization and the nutritional composition and microbiota differ.^93^ Additionally, ∼93% of infants with SV CHD are prescribed a high–calorie diet at S1P discharge, often by adding infant formula or bovine-milk-derived fortifier to HM.^12^ It is unclear whether an infant fed only HM plus fortification was considered to receive exclusive HM, and it is possible that clinicians at different sites defined exclusive HM differently. Definitions of outcomes (eg, NEC) could also have varied across sites. Analyses of the preoperative time only included the 57.8% of infants who were enterally fed, and the results may not be generalizable to all infants with SV CHD.

Considering the known association between HM/BF and maternal factors such as race and/or economic status, our inclusion of variables derived from an infant’s ZCTA are a strength of this study. However, these variables only approximate an individual family’s situation. Moreover, we did not have information on maternal intent for HM/BF or factors such as previous BF experience that could impact self-efficacy for these feeding practices. Future multisite, prospective, longitudinal studies with careful measurement of the volume and dose of HM/BF throughout the infant’s first year of life should also include detailed analysis of relevant family, maternal, and social factors.

### Conclusion

In this large, multisite study using robust statistical techniques designed to reduce bias and support causal inference, we found that infants with high HM feeding and direct BF exposures experienced multiple significant improvements in outcomes for infants with SV CHD, including reduced incidence of NEC, infection, and sepsis; substantially shorter length of stay at both S1P and S2P surgeries; and lower mortality. These results align with previous research including preterm and other surgical neonates; however, HM/BF research in the context of CHD currently lags behind that focused on preterm populations. Future work is urgently needed to confirm the results of this study and to identify potential mechanistic causal pathways for improved outcomes. Most importantly, this study highlights the critical need for improved, condition-specific lactation support to address the currently low rates of HM and BF for infants with SV CHD. Our findings demonstrate that increasing the dose and duration of HM and direct BF has strong potential to substantially improve the health outcomes of these vulnerable infants.

## Data Availability

The data are held by the National Pediatric Cardiology Quality Improvement Project and are not publicly available.

## Acknowledgements

The authors would like to thank the NPC-QIC for providing the data for this study, and also Barbara McMorris, PhD, for providing helpful feedback in refining the study design.

## Supplemental Material

Figures S1–S3

Table S1

