## Supplemental Material for "Human milk feeding and direct breastfeeding improve outcomes for infants with single ventricle congenital heart disease: Propensity score matched analysis of the NPC-QIC registry"

**Figure S1.** Flow diagram for inclusion and exclusion at all study time points

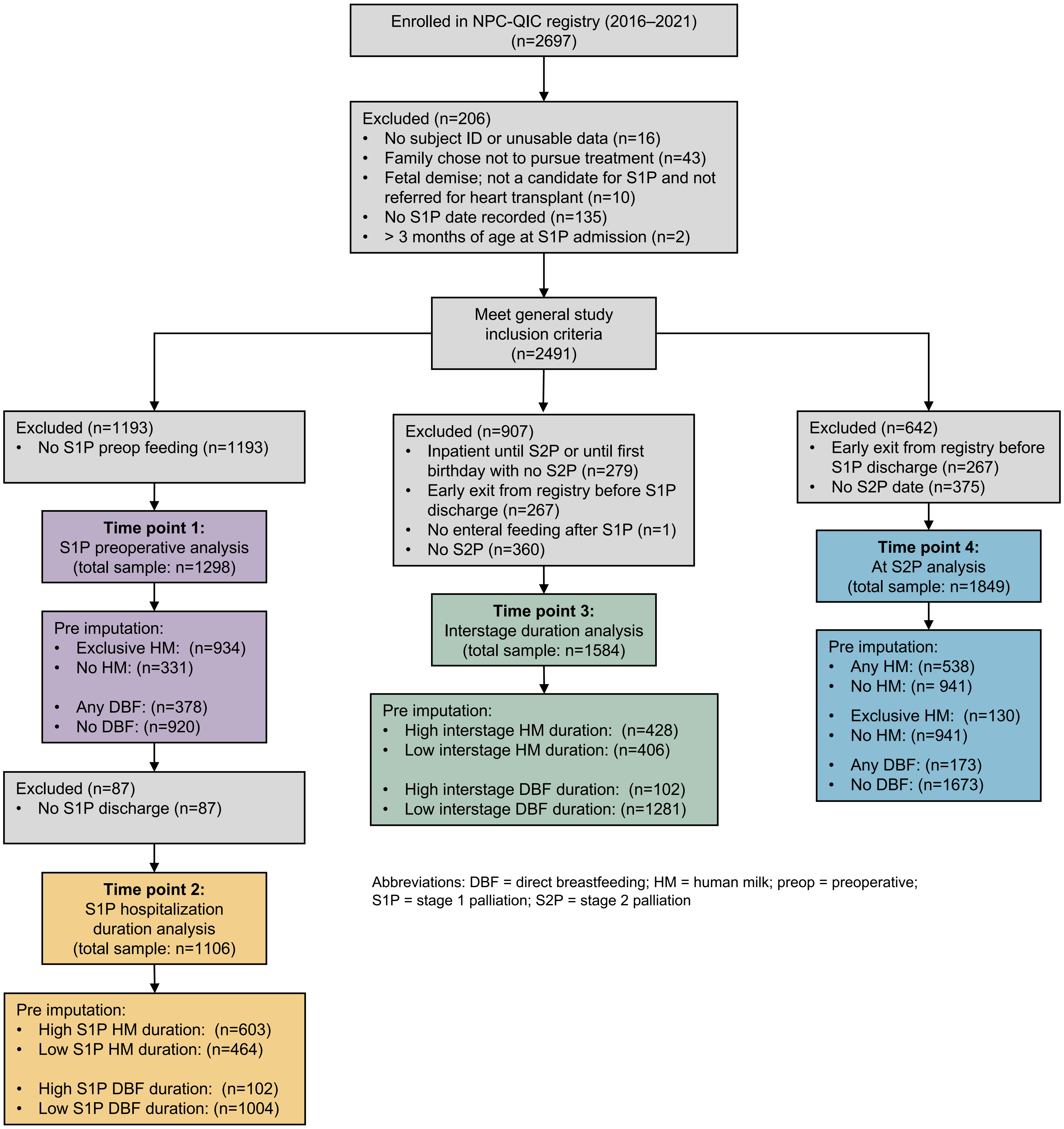


Abbreviations: DBF = direct breastfeeding; HM = human milk; preop = preoperative; S1P = stage 1 palliation; S2P = stage 2 palliation

**Figure S2.** Visualization of standardized mean differences indicating covariate balance in the original cohort and after propensity score matching for exposures and outcomes examined during the stage 1 palliation hospitalization

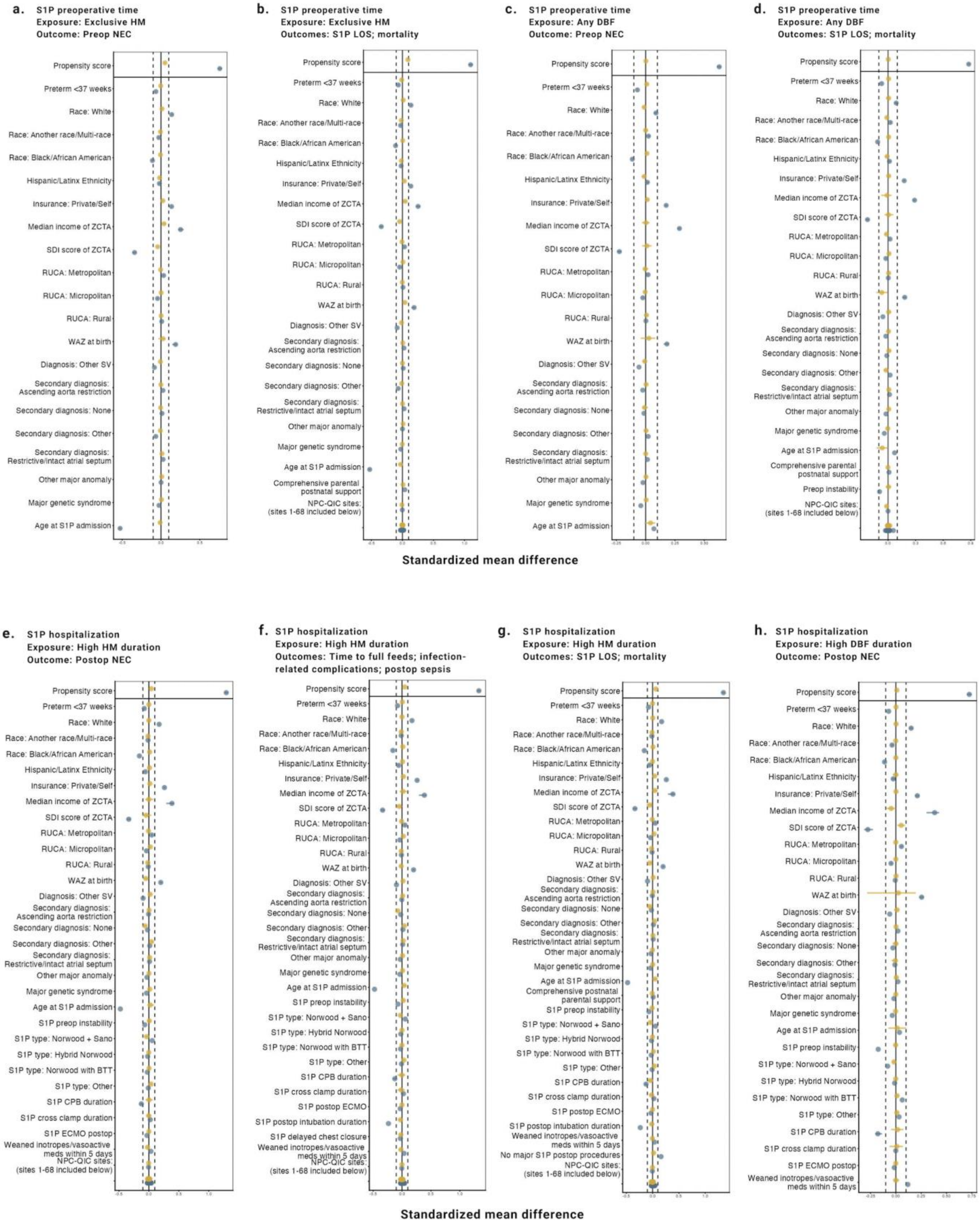

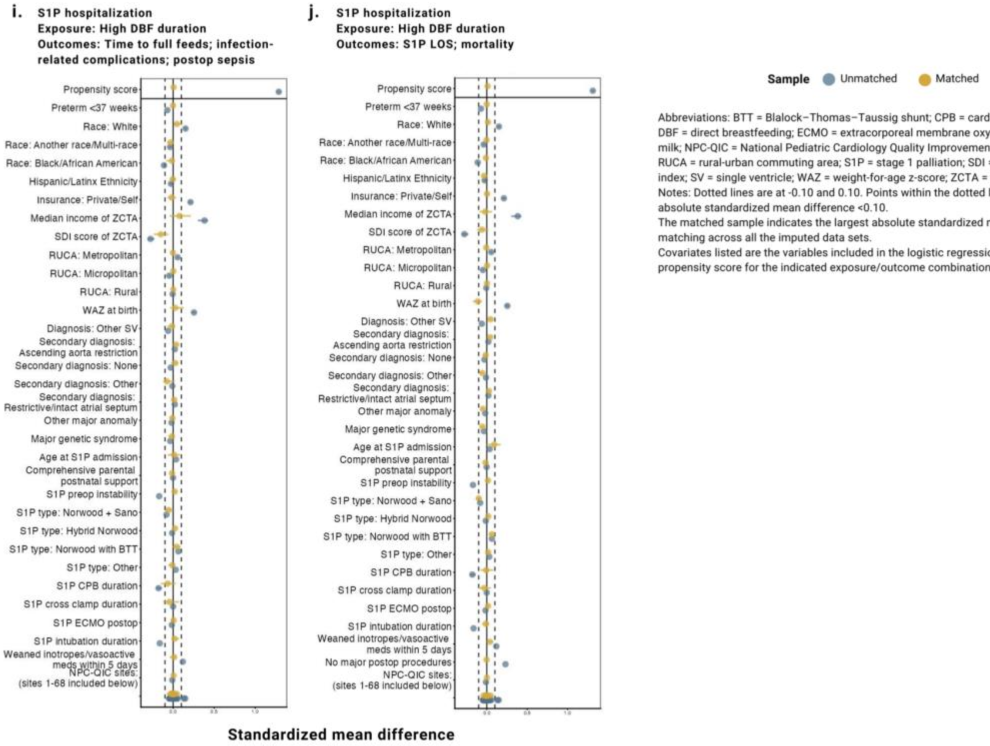


Abbreviations: BTT = Blalock–Thomas–Taussig shunt; CPB = cardiopulmonary bypass; DBF = direct breastfeeding; ECMO = extracorporeal membrane oxygenation; HM = human milk; NPC-QIC = National Pediatric Cardiology Quality Improvement Collaborative; RUCA = rural-urban commuting area; S1P = stage 1 palliation; SDI = social deprivation index; SV = single ventricle; WAZ = weight-for-age z-score; ZCTA = zip code tabulation area

Notes: Dotted lines are at -0.10 and 0.10. Points within the dotted lines indicate an absolute standardized mean difference <0.10.

The matched sample indicates the largest absolute standardized mean difference after matching across all the imputed data sets.

Covariates listed are the variables included in the logistic regression model to create the propensity score for the indicated exposure/outcome combination.

**Figure S3.** Visualization of standardized mean differences indicating covariate balance in the original cohort and after propensity score matching for exposures and outcomes examined during the interstage period and at stage 2 palliation

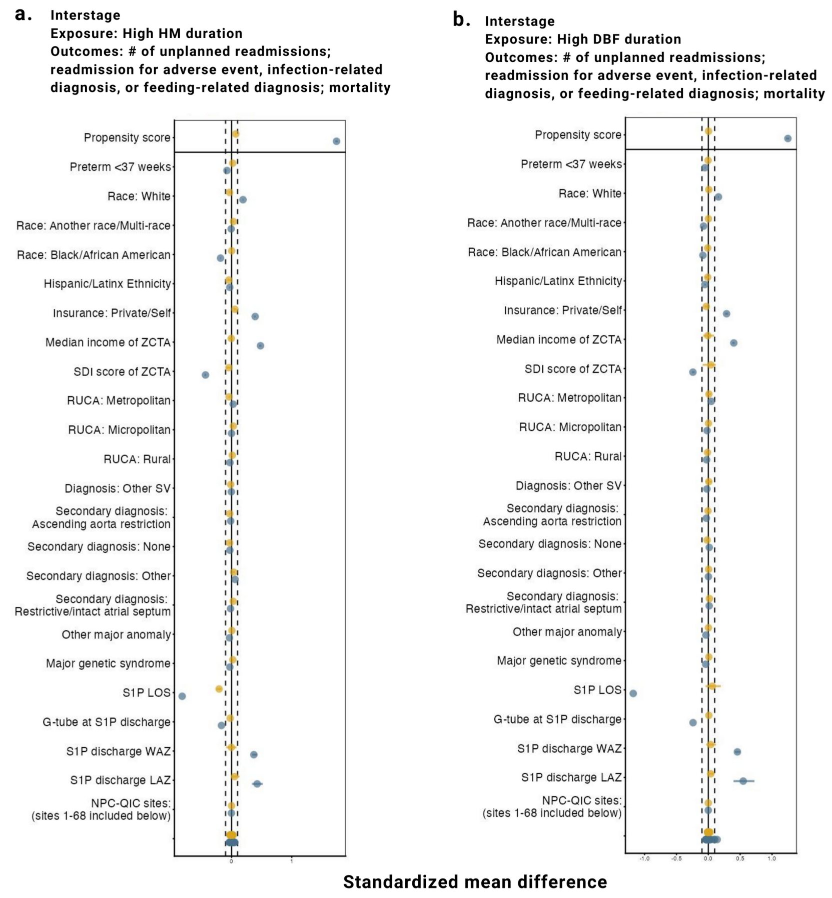


**Sample** Unmatched Matched

Abbreviations: DBF = direct breastfeeding; G-tube = gastrostomy tube; HM = human milk feeding; LAZ = length-for-age z-score; NPC-QIC = National Pediatric Cardiology Quality Improvement Collaborative; RUCA = rural-urban commuting area; S1P = stage 1 palliation; S2P = stage 2 palliation; SDI = social deprivation index; SV = single ventricle; WAZ = weight-for-age z-score; ZCTA = zip code tabulation area

Notes: Dotted lines are at -0.10 and 0.10. Points within the dotted lines indicate an absolute standardized mean difference <0.10. The matched sample indicates the largest absolute standardized mean difference after matching across all the imputed data sets. Covariates listed are the variables included in the logistic regression model to create the propensity score for the indicated exposure/outcome combination.

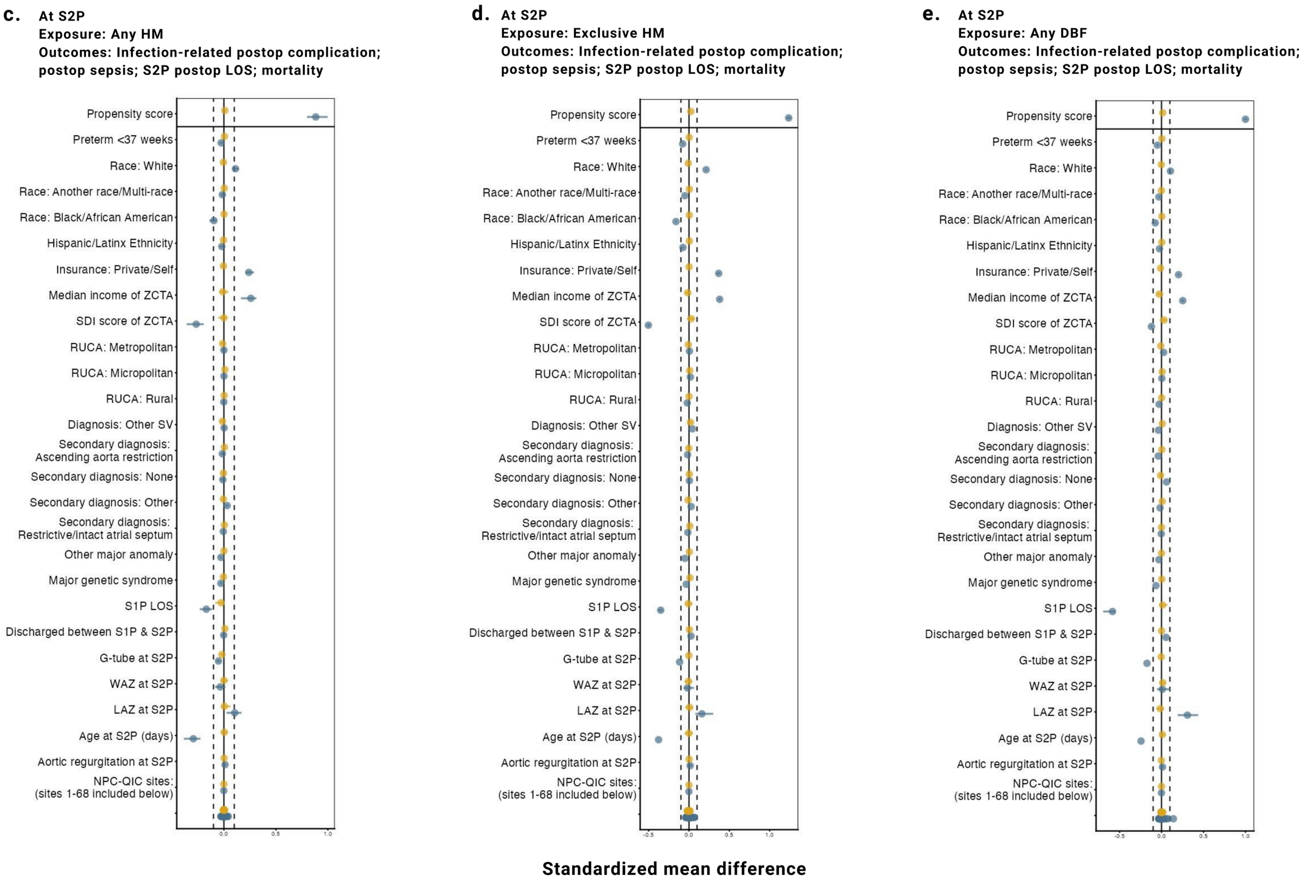

**Table S1.** Details of propensity score development for feeding exposure/outcome combinations at time points from the stage 1 palliation preoperative time to stage 2 palliation

| | Sample size: Original cohort; Exposed/Control | Average n (%) of unmatched exposed across imputations | Average n (%) of unmatched controls across imputations | Caliper | k:1 ratio | Max # of controls reused | Variables included in all propensity score models / Additional variables added to propensity score models for specific outcomes | Unbalanced variables added to outcome regression model |
|---|---|---|---|---|---|---|---|---|
| **S1P preoperative time** | | | | | | | | |
| Exclusive HM | N = 1265; 934/331 | | | | | | preterm, race, Hispanic/Latino/a ethnicity, insurance type, median income of ZCTA, SDI score of ZCTA, RUCA of ZCTA, age at S1P admission, WAZ at birth, primary & secondary cardiac diagnoses, other major anomalies, major genetic syndrome | |
| Preop NEC | | 1.95 (0.2) | 11.3 (3.4) | 0.2 | 4 | 10 | N/A | N/A |
| S1P LOS | | 66.35 (7.1) | 17.6 (5.3) | 0.2 | 4 | 8 | / clinical site, comprehensive postnatal parental support | N/A |
| All-cause mortality | | 66.35 (7.1) | 17.6 (5.3) | 0.2 | 4 | 8 | / clinical site, comprehensive postnatal parental support | N/A |
| Any direct BF | N = 1298; 378/920 | | | | | | | |
| Preop NEC | | 0.25 (0.1) | 287.25 (31.2) | 0.2 | 4 | 5 | N/A | N/A |
| S1P LOS | | 5.65 (1.5) | 367.75 (40.0) | 0.2 | 4 | 15 | / clinical site, comprehensive postnatal parental support, S1P preoperative instability | age at S1P admission, WAZ at birth |
| All-cause mortality | | 5.65 (1.5) | 367.75 (40.0) | 0.2 | 4 | 15 | / clinical site, comprehensive postnatal parental support, S1P preoperative instability | age at S1P admission, WAZ at birth |
| **During the S1P hospitalization (preoperative – discharge)** | | | | | | | | |
| Exclusive HM preop & any HM at discharge vs. Any type of preop feeding & no HM at discharge | N = 1067; 603/464 | | | | | | preterm, race, Hispanic/Latino/a ethnicity, insurance type, median income of ZCTA, SDI score of ZCTA, RUCA of ZCTA, primary & secondary cardiac diagnoses, other major anomalies, major genetic syndrome, age at S1P admission, WAZ at birth, S1P preoperative instability, S1P surgical procedure, weaned inotropes and vasoactive meds within 5 days post-S1P, S1P CPB duration, S1P cross clamp duration, need for ECMO post-S1P, clinical site | |
| Postop NEC | | 6.95 (1.2) | 77.5 (16.7) | 0.2 | 4 | 20 | N/A | SDI score of ZCTA |
| Infection-related postop complication (sepsis, pneumonia, wound infection) | | 7.3 (1.2) | 85.4 (18.4) | 0.2 | 4 | 20 | / delayed sternal closure post-S1P, intubation duration post-S1P | N/A |
| Postop sepsis | | 7.3 (1.2) | 85.4 (18.4) | 0.2 | 4 | 20 | / delayed sternal closure post-S1P, intubation duration post-S1P | N/A |
| Time to full feeds | | 7.3 (1.2) | 85.4 (18.4) | 0.2 | 4 | 20 | / delayed sternal closure post-S1P, intubation duration post-S1P | N/A |
| S1P LOS | | 6.8 (1.1) | 87.75 (18.9) | 0.2 | 4 | 15 | / intubation duration post-S1P, any major S1P postop procedures, comprehensive postnatal parental support | SDI score of ZCTA |
| All-cause mortality | | 6.8 (1.1) | 87.75 (18.9) | 0.2 | 4 | 15 | / intubation duration post-S1P, any major S1P postop procedures, comprehensive postnatal parental support | SDI score of ZCTA |
| Any preop direct BF & any direct BF at discharge vs. Any type of preop feeding & no direct BF at S1P discharge | N = 1106; 102/1004 | | | | | | | |
| Postop NEC | | 2.7 (2.7) | 464.5 (46.3) | 0.2 | 10 | 2 | N/A | age at S1P admission, WAZ at birth |
| Infection-related postop complication (sepsis, pneumonia, wound infection) | | 2.95 (3.0) | 730.6 (72.8) | 0.3 | 10 | 10 | / delayed sternal closure post-S1P, intubation duration post-S1P | age at S1P admission, WAZ at birth, S1P CPB duration, S1P cross clamp duration, S1P surgical procedure, SDI score of ZCTA, median income of ZCTA |
| Postop sepsis | | 1.35 (1.3) | 741.45 (73.9) | 0.3 | 10 | 10 | / delayed sternal closure post-S1P, intubation duration post-S1P | age at S1P admission, WAZ at birth, S1P cross clamp duration, S1P surgical procedure, SDI score of ZCTA |
| Time to full feeds | | 1.35 (1.3) | 741.45 (73.9) | 0.3 | 10 | 10 | / delayed sternal closure post-S1P, intubation duration post-S1P | age at S1P admission, WAZ at birth, S1P CPB duration, S1P cross clamp duration, S1P surgical procedure, SDI score of ZCTA, median income of ZCTA |
| S1P LOS | | 1.35 (1.3) | 741.45 (73.9) | 0.3 | 10 | 10 | / intubation duration post-S1P, any major S1P postop procedures, comprehensive postnatal parental support | age at S1P admission, WAZ at birth, S1P cross clamp duration, S1P surgical procedure, SDI score of ZCTA |
| All-cause mortality | | 1.35 (1.3) | 741.45 (73.9) | 0.3 | 10 | 10 | / intubation duration post-S1P, any major S1P postop procedures, comprehensive postnatal parental support | age at S1P admission, WAZ at birth, S1P cross clamp duration, S1P surgical procedure, SDI score of ZCTA |

| | | | | | | | | |
|---|---|---|---|---|---|---|---|---|
| **During the interstage period (S1P discharge – S2P)** | | | | | | | | |
| Any HM at S1P discharge & any HM at S2P vs. No HM at S1P discharge & no HM at S2P | N = 836; 428/408 | | | | | | preterm, race, Hispanic/Latinx ethnicity, insurance type, median income of ZCTA, SDI score of ZCTA, RUCA of ZCTA, primary & secondary cardiac diagnoses, other major anomalies, major genetic syndrome, S1P hospital LOS, G-tube at S1P discharge, S1P discharge WAZ, S1P discharge LAZ, clinical site | |
| # of unplanned interstage readmissions | | 0 (0.0) | 96.4 (23.6) | 0.2 | 8 | 20 | N/A | S1P discharge LAZ, S1P hospital LOS |
| Any interstage readmission for adverse event | | | | (same for all outcomes) | | | N/A | (same for all outcomes) |
| Infection-related interstage readmission | | | | (same for all outcomes) | | | N/A | (same for all outcomes) |
| Feeding-related interstage readmission | | | | (same for all outcomes) | | | N/A | (same for all outcomes) |
| All-cause mortality | | | | (same for all outcomes) | | | N/A | (same for all outcomes) |
| Any direct BF at S1P discharge & any direct BF at S2P vs. No direct BF at S1P discharge & no direct BF at S2P | N = 1383; 102/1281 | | | | | | | |
| # of unplanned interstage readmissions | | 2.9 (2.8) | 989.35 (70.6) | 0.2 | 10 | 10 | N/A | S1P discharge WAZ, SDI score of ZCTA, S1P hospital LOS |
| Any interstage readmission for adverse event | | | | (same for all outcomes) | | | N/A | (same for all outcomes) |
| Infection-related interstage readmission | | | | (same for all outcomes) | | | N/A | (same for all outcomes) |
| Feeding-related interstage readmission | | | | (same for all outcomes) | | | N/A | (same for all outcomes) |
| All-cause mortality | | | | (same for all outcomes) | | | N/A | (same for all outcomes) |
| **At S2P** | | | | | | | | |
| Any HM vs. No HM | N = 1847; 785/1062 | | | | | | preterm, race, Hispanic/Latinx ethnicity, insurance type, median income of ZCTA, SDI score of ZCTA, RUCA of ZCTA, primary & secondary cardiac diagnoses, other major anomalies, major genetic syndrome, S1P hospital LOS, G-tube at S2P, WAZ at S2P, LAZ at S2P, age at S2P, aortic regurgitation at S2P, discharged between S1P and S2P, clinical site | |
| S2P postop LOS; | | 2.85 (0.0) | 243.3 (22.9) | 0.2 | 4 | 10 | N/A | N/A |
| Infection-related S2P postop complication | | | | (same for all outcomes) | | | N/A | N/A |
| S2P postop sepsis | | | | (same for all outcomes) | | | N/A | N/A |
| All-cause mortality | | | | (same for all outcomes) | | | N/A | N/A |
| Exclusive HM vs. No HM | N = 1077; 130/947 | | | | | | | |
| S2P postop LOS | | 1.35 (1.0) | 533.8 (56.4) | 0.2 | 10 | 2 | N/A | N/A |
| Infection-related S2P postop complication | | | | (same for all outcomes) | | | N/A | N/A |
| S2P postop sepsis | | | | (same for all outcomes) | | | N/A | N/A |
| All-cause mortality | | | | (same for all outcomes) | | | N/A | N/A |
| Any direct BF vs. No direct BF | N = 1847; 173/1674 | | | | | | | |
| S2P postop LOS | | 4.1 (2.4) | 1009.2 (60.3) | 0.2 | 10 | 2 | N/A | N/A |
| Infection-related S2P postop complication | | | | (same for all outcomes) | | | N/A | N/A |
| S2P postop sepsis | | | | (same for all outcomes) | | | N/A | N/A |
| All-cause mortality | | | | (same for all outcomes) | | | N/A | N/A |

Abbreviations: BF = breastfeeding; CPB = cardiopulmonary bypass; ECMO = extracorporeal membrane oxygenation; G-tube = gastrostomy; HM = human milk; LAZ = length-for-age z-score; LOS = length of stay; RUCA = rural-urban commuting area; S1P = stage 1 palliation; S2P = stage 2 palliation; SDI = social deprivation index; WAZ = weight-for-age z-score; ZCTA = zip code tabulation area
